# Pharmacokinetics and predicted neutralization coverage of VRC01 in HIV-uninfected participants of the Antibody Mediated Prevention (AMP) trials

**DOI:** 10.1101/2020.09.02.20182881

**Authors:** Yunda Huang, Logashvari Naidoo, Lily Zhang, Lindsay N. Carpp, Erika Rudnicki, April Randhawa, Pedro Gonzales, Adrian McDermott, Julie Ledgerwood, Margarita M. Gomez Lorenzo, David Burns, Allan DeCamp, Michal Juraska, John Mascola, Srilatha Edupuganti, Nyaradzo Mgodi, Myron Cohen, Larry Corey, Philip Andrew, Shelly Karuna, Peter B. Gilbert, Kathryn Mngadi, Erica Lazarus

## Abstract

The phase 2b AMP trials are testing whether the broadly neutralizing antibody VRC01 prevents HIV-1 infection in two cohorts: women in sub-Saharan Africa, and men and transgender persons who have sex with men (MSM/TG) in the Americas and Switzerland. We used nonlinear mixed effects modeling of longitudinal serum VRC01 concentrations to characterize pharmacokinetics and predict HIV-1 neutralization coverage. We found that body weight significantly influenced clearance, and that the mean peripheral volume of distribution, steady state volume of distribution, elimination half-life, and accumulation ratio were significantly higher in MSM/TG than in women. Neutralization coverage was predicted to be higher in the first (versus second) half of a given 8-week infusion interval, and appeared to be higher in MSM/TG than in women overall. Study cohort differences in pharmacokinetics and neutralization coverage provide insights for interpreting the AMP results and for investigating how VRC01 concentration and neutralization correlate with HIV incidence.

## Introduction

With an estimated 37.9 million people living with HIV and 770,000 deaths due to AIDS-related causes in 2018 [1], the global HIV pandemic continues to deal a devastating blow to public health. Advances such as antiretroviral therapy and pre-exposure prophylaxis (PrEP) have significantly reduced AIDS-related morbidity and mortality and HIV acquisition, but challenges in access, uptake and adherence continue [2]. In addition, the rollout of PrEP has had a variable effect on HIV acquisition, particularly in the absence of a supporting comprehensive combination prevention program [3]. An international commission of global experts and stakeholders recently concluded that “existing HIV tools and strategies are insufficient” to end the HIV pandemic [4], highlighting the need for new and complementary preventive interventions.

Monoclonal broadly neutralizing antibodies (bnAbs) against HIV-1 are a promising new avenue for HIV-1 prevention [5]. VRC01 is a human IgG1 monoclonal bnAb that targets the conserved CD4 binding site on the HIV-1 envelope (Env) surface glycoprotein [6], demonstrates breadth of neutralization of clinical HIV-1 isolates [7, 8], and prevents simian HIV infection in nonhuman primates [9-15]. In addition, it has been shown to be safe and well-tolerated in phase 1 trials in healthy HIV-uninfected adults at low-risk of HIV-1 acquisition in the United States when administered subcutaneously or intravenously (IV) in 4-weekly to 8-weekly doses [16, 17]. Population pharmacokinetic (PK) modeling of these trials [18] demonstrated that following intravenous administration, VRC01 PK was best described by an open 2-compartment disposition model with first-order elimination from the central compartment, which accounts for reversible monoclonal antibody (mAb) transfer between the central and peripheral compartments [19]. VRC01 half-life estimates were consistent between the two phase 1 clinical trials conducted in the US [16, 17] and VRC01 PK features were relatively stable across the multiple doses [17]. The pharmacokinetics of mAbs is such that biodistribution is mainly in the vascular and interstitial spaces [20], and is dependent on extravasation into tissue spaces, distribution in the interstitial fluid, mAb binding to tissue components, and clearance from the tissues [19]. Moreover, due to their relatively large size (molecular weight approximately 150 kDa), mAbs cannot be eliminated from the kidneys and are instead eliminated mainly through intracellular proteolytic catabolism by lysosomes to amino acids and smaller peptides that are then reused for new protein synthesis [19, 20].

VRC01 is the first bnAb being tested for efficacy for the prevention of HIV-1 infection in humans in the proof-of-concept Antibody Mediated Prevention (AMP) trials [21], with primary results expected in Q4 2020. The safety and efficacy of ten 8-weekly IV infusions of VRC01 are being assessed in AMP in two study populations at risk of HIV-1 acquisition through predominantly different transmission routes: women in sub-Saharan Africa who have sex with men (HVTN 703/HPTN 081; ClinicalTrials.gov #NCT02568215) and men and transgender persons in Brazil, Peru, Switzerland, and the United States who have sex with men (MSM/TG) (HVTN 704/HPTN 085; ClinicalTrials.gov #NCT02716675) [21]. The major HIV-1 subtypes also differ between the two trials, with clade C predominating in sub-Saharan Africa and clade B predominating in the Americas and Switzerland [22].

One key secondary objective of AMP is to assess, through a case-control study, marker correlates (or predictors) of instantaneous HIV-1 risk (see [21] and [23] for further details), e.g. VRC01 serum concentration and serum neutralization titer to panels of HIV-1 isolates. If validated, such a concentration or neutralization biomarker will aid HIV vaccine development by setting a benchmark biomarker value for the required potency of a vaccine-induced neutralizing antibody response to putatively achieve a high level of protection against HIV infection. This could help define study endpoints in phase 1 and 2 trials that vet candidate HIV vaccines for advancement into efficacy trials.

In preparation for the AMP case-control correlates study, we conducted a PK pilot study among a subset of VRC01 recipients in the AMP trials who remained HIV-uninfected until the end of the study and were not taking PrEP during the study. The objectives were to develop a population PK (popPK) model to characterize the PK features of VRC01 in more diverse HIV risk settings and populations than the phase 1 trials, over 10 administrations rather than the 3-4 administrations previously considered, and to identify factors that may influence these PK features. Consequently, this popPK pilot study provides a technique for simulating serum concentrations for all VRC01 recipients and for inferring neutralization coverage of participants’ sera against the circulating strains in the AMP trials. A similar technique will be used to estimate VRC01 concentrations for individual participants in the case-control cohort at any given day during follow-up, which constitutes a critical data component in the AMP case-control correlates study. Importantly, findings from this PK pilot study are expected to aid the interpretation of the final AMP trial results on prevention efficacy, and to inform the sampling design of the case-control correlates study.

## Results

### Study population descriptions

The characteristics of the 47 AMP pilot PK study participants are summarized in Table 1. Overall, 24 of the 47 participants were assigned female sex at birth, 28 were black, and the median age was 26 (range 19 to 50). In HVTN 703/HPTN 081, all 23 participants were assigned female sex at birth, 22 were black, and the median age was 25 years (range 19 to 37). In HVTN 704/HPTN 085, 23 of the 24 participants were assigned male sex at birth, 6 were black, and the median age was 31 years (range 19 to 50).

**Table 1:**
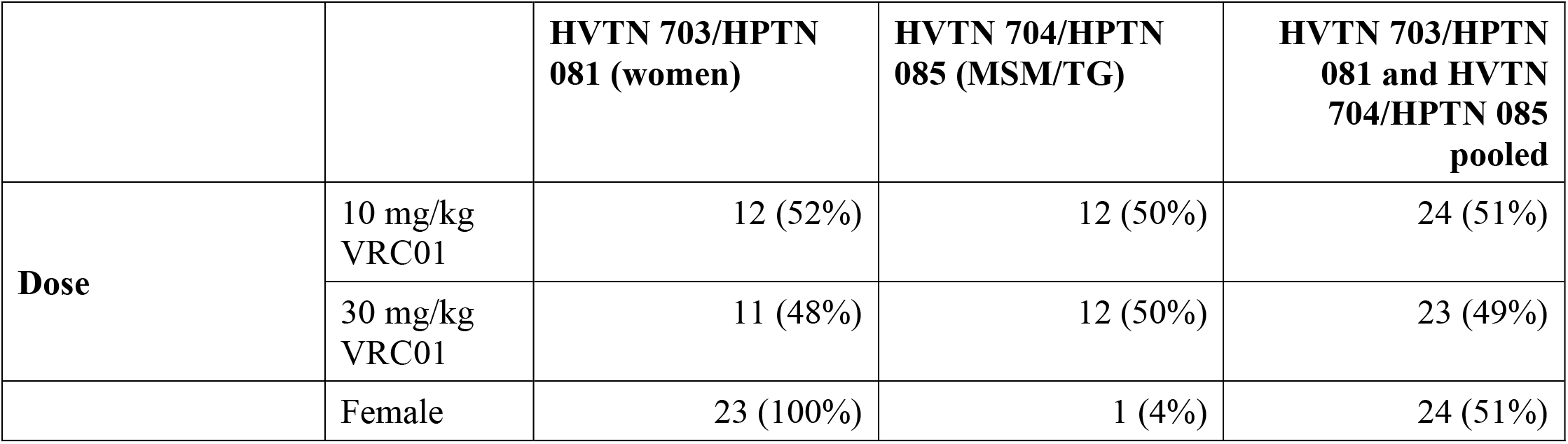
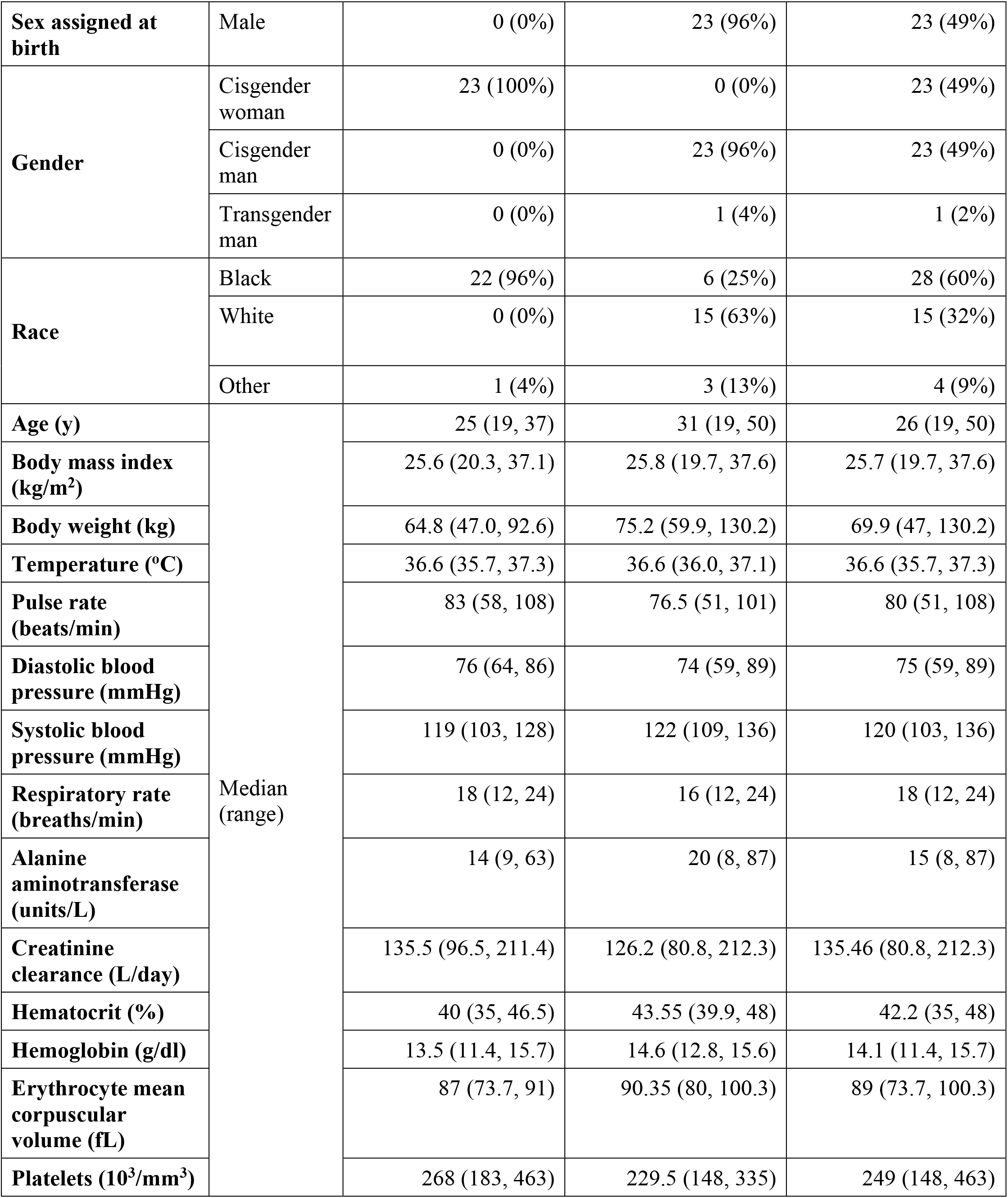
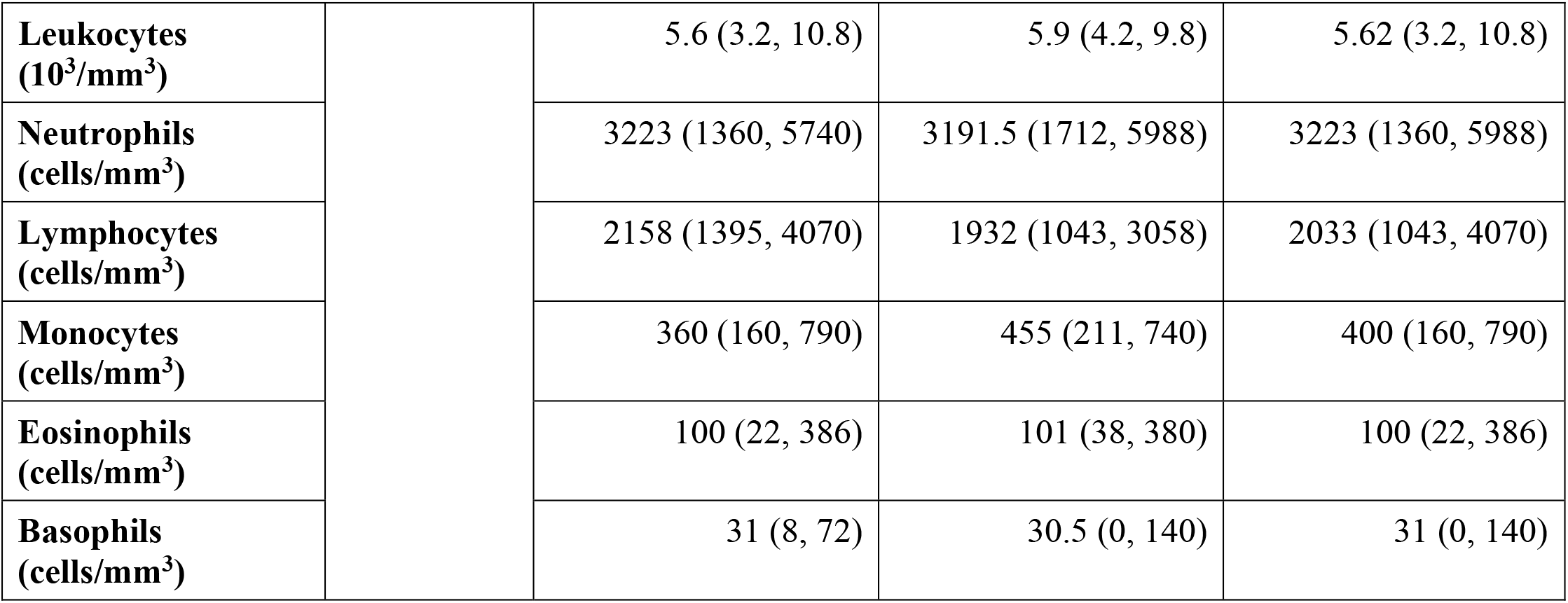
Characteristics of AMP participants at enrollment included in the PK pilot study.

Forty-five (95.7%) of the 47 pilot study participants received the planned 10 infusions of VRC01. In HVTN 703/HPTN 081 (women), the average interval between two consecutive infusions was 59.7 days (range 48.7 to 103) with one participant in the 30 mg/kg dose group who missed all infusions starting from the 4^th^; in HVTN 704/HPTN 085 (MSM/TG), the average infusion interval was 57.7 days (range 48.1 to 106) with one participant in the 30 mg/kg dose group who missed the 9^th^ infusion (Figure S1). Given that baseline serum concentration measurements were all negative at week 0 (serving as assay quality control), and only one participant had a detectable level at week 96 (Figure S2), the popPK modeling excluded data at weeks 0, 96 and 104, and included 1003 VRC01 serum concentrations between week 4 through 88 from the 47 participants (Figure 1).

**Figure 1:**
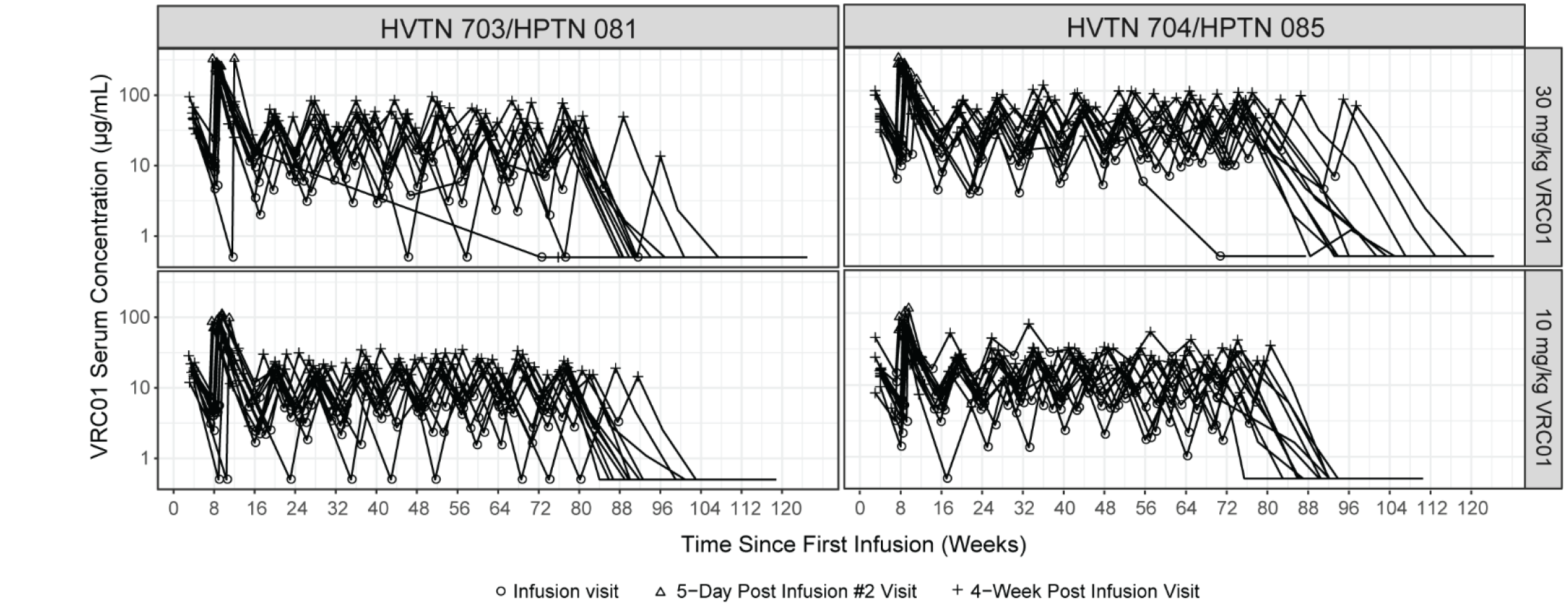
Individual-level VRC01 serum concentration (log10-scale) over time. A) Data from HVTN 703/HPTN 081 (women) (n=23), B) Data from HVTN 704/HPTN 085 (MSM/TG) (n=24). “+” indicates the observed concentration at a 4-week post infusion visit, an open circle indicates the observed concentration at an infusion visit, and a triangle indicates the observed 143 concentration at the 5-day post infusion #2 visit.

### Base popPK models

Dosing at 10 mg/kg or 30 mg/kg did not influence any of the PK parameters as dose is evaluated as a covariate in the popPK models, thereby verifying the linear PK assumption regarding PK parameters having the same value across dose levels and allowing a single PK model to be used to describe data for both dose groups. In the base two-compartment model, the combination proportional + additive error model was chosen for its significantly improved objective function value (OFV=3156) over the proportional (OFV=3313) and additive (OFV=4373) models. IIV was observed primarily for CL and Vp with %CV of 20-21% and 24-26%, respectively, with and without IOV being considered (Table S1). However, after IOV was considered, residual errors were considerably reduced, and the precision of the fixed effects and the constant error term were considerably improved (Table S1). Therefore, PK features including CL, Vp, half-life, steady state AUC and accumulation ratio were estimated from the base model with IOV considered. Of note, despite the considerable level of IOV, the individual-level CL, Vp and half-life estimates did not obviously increase or decrease as the infusion numbers progressed (Figure 2).

**Figure 2:**
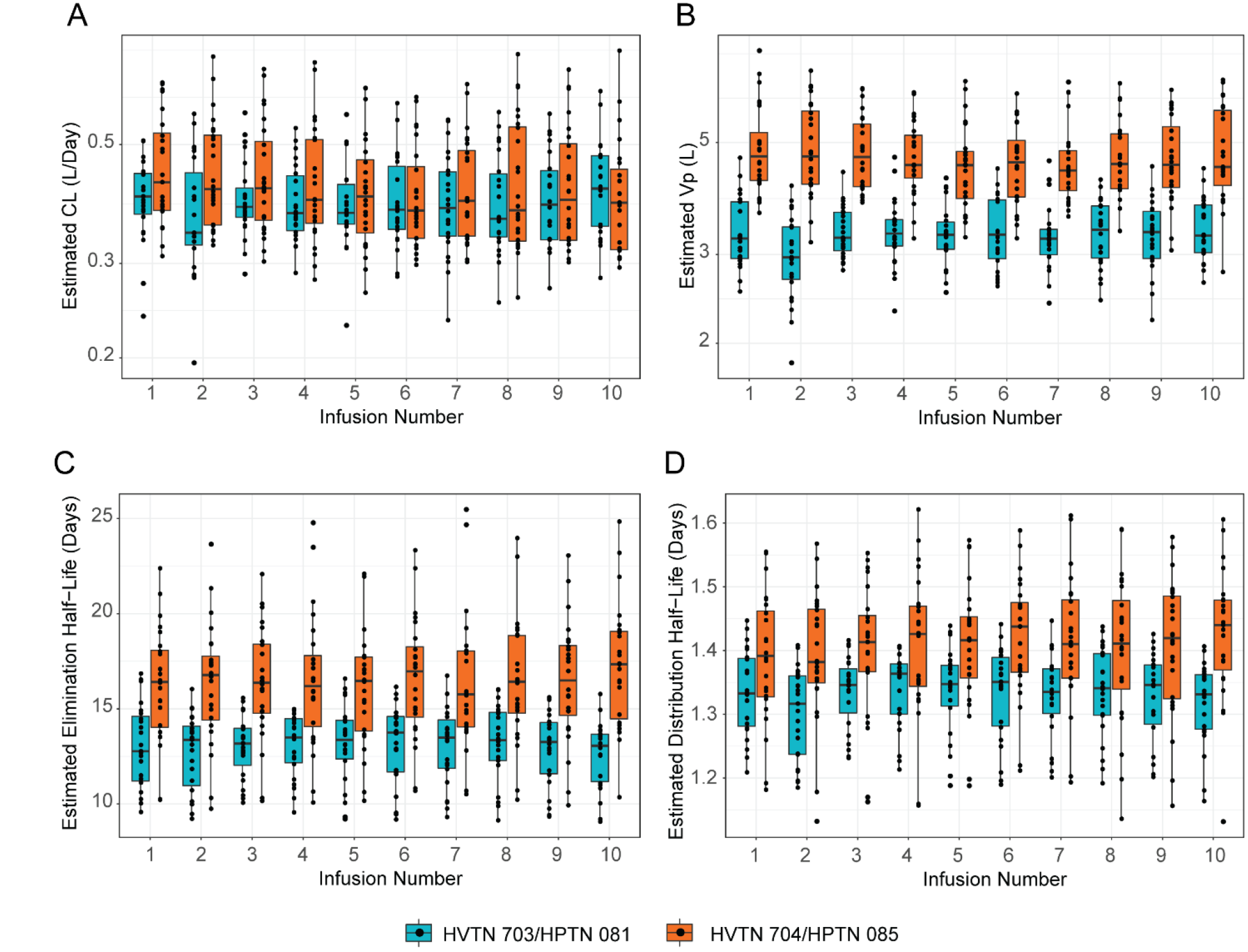
Distributions of individual-level PK parameter estimates of VRC01 over the 10 infusions. A) Clearance (CL), and B) volume of the peripheral compartment (Vp), C) distribution half-life, and D) elimination half-life estimates are shown. Estimates are based on the inter-occasion variability-included base model described in Table S1. Green and purple points and curves are used for HVTN 703/HPTN 081(women) and HVTN 704/HPTN 085(MSM/TG), respectively. Panels A and B are shown on a log10 scale since both CL and V_p_ show a log-normal distribution, while panels C and D are shown on a linear scale.

### Final popPK models

Clinical and demographic variables were assessed for their potential role in explaining the observed inter-individual variability according to the model selection process described. Further details on the construction of the final popPK model are given in Text S1. As shown in Table S2, when IOV was included, the model fit significantly improved (OFV = - 1434.42 vs. OFV = 3064.36), suggesting that IOV will likely need to be considered in the estimation of concentrations for the AMP case-control correlates study. Overall, the popPK model diagnostic results suggested that the modeling assumptions were reasonable, and the final model with IOV included provides a reliable description of the data (Figures S5 and S6).

Based on the final popPK model, the population mean estimate for CL was 0.383 (95% CI: 0.357, 0.409) L/day for individuals with a body weight of 68.8 kg (median body weight over both studies), with an estimated 0.611 (95% CI: 0.350, 0.872) log increase of CL per kg of body weight. The population mean estimate for Vp was 3.17 L (95% CI: 2.87, 3.47) for individuals in HVTN 703/HPTN 081 (MSM/TG), estimated to be 0.428 (95% CI: 0.270, 0.586) fold higher for individuals in HVTN 704/HPTN 085 (women). After accounting for body weight and study cohort, the inter-individual variability of CL and Vp decreased from 20.0 to 15.8 %CV, and from 23.6 to 13.5 %CV, respectively.

In addition, the terminal half-life of VRC01estimated to be 12.33 and 16.43 days, distribution half-life 1.24 and 1.33 days, and steady state volume of distribution 5.26 and 6.62 L in HVTN 703/HPTN 081 (women) and HVTN 704/HPTN 085 (MSM/TG), respectively, based on the final popPK model. In the 10mg/kg dose group, the final model resulted in an estimated accumulation ratio of 1.04 and 1.09 for the average 60 days of dosing interval in the study, and steady state AUC of 1796.35 and 1796.35 mg*day/mL in HVTN 703/HPTN 081 (women) and HVTN 704/HPTN 085 (MSM/TG), respectively, for individuals with a body weight of 68.8 kg. In the 30 mg/kg dose group, the final model resulted in an estimated accumulation ratio of 1.04 and 1.09, and steady state AUC of 5389.03 and 5389.03 mg*day/mL in HVTN 703/HPTN 081 (MSM/TG) and HVTN 704/HPTN 085 (women), respectively.

### Predicted neutralization coverage

To visualize the expected concentrations in the two AMP trials, Figure 3 and Figure S7 display simulated concentrations over 8 weeks after a single dose for the two dose groups. These concentrations were simulated based on the final popPK model accounting for body weight of actual AMP trial participants. Figure 3 also displays the predicted coverage of VRC01 after a single dose based on known IC50 values in each trial population. The predicted coverage of VRC01 appears to be higher in HVTN 704/HPTN 085 (MSM/TG) compared to HVTN 703/HPTN 081 (women), for both dose groups after a given dose (Figures 3 and S7) and over the course of 10 doses (Figure S8). As expected, coverage is predicted to be considerably higher in the first 4 weeks post-infusion compared to in the second 4 weeks [for 10 mg/kg groups: 43% vs 14% in HVTN 704/HPTN 085 (MSM/TG), 34% vs 7% in HVTN 703/HPTN 081 (women); for 30 mg/kg groups: 68% vs 35% in HVTN 704/HPTN 085 (MSM/TG), 56% vs 22% in HVTN 703/HPTN 081 (women)].

**Figure 3:**
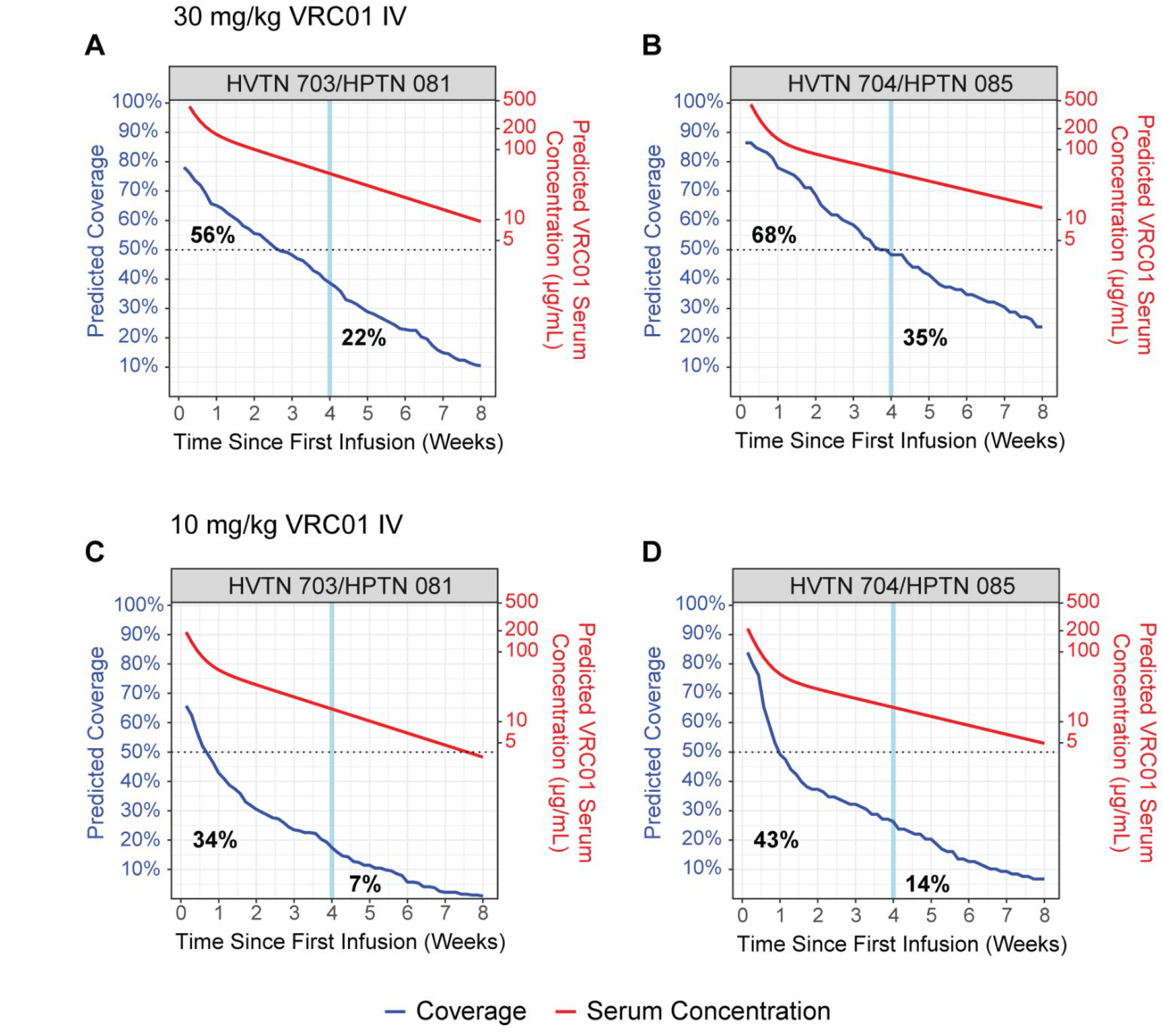
Predicted VRC01 neutralization coverage and serum concentration by time since first infusion. A (30 mg/kg), C (10 mg/kg) in HVTN 703/HPTN 081 (women): Percent of 315 clade C isolates on CATNAP that would be sensitive to VRC01 neutralization if the geometric mean serum concentration at the given time-point was at least 100-fold greater than the viral in vitro inhibitory concentration 50% (IC50). B (30 mg/kg), D (10 mg/kg) in HVTN 704/HPTN 085 (MSM/TG): Percent of 118 clade B isolates on CATNAP that would be sensitive to VRC01 neutralization if the geometric mean serum concentration at the given time-point was at least 100-fold greater than the viral in vitro inhibitory concentration 50% (IC50). Within each plot, the left-most bolded percentage corresponds to the average coverage in the first 4 weeks post-first infusion and the right-most bolded percentage corresponds to the average coverage in the second 4 weeks post-first infusion.

### Covariate-adjusted study effects on PK features estimated from the base popPK model and TMLE

Seven individual-level PK features: CL, Vp, dose-normalized steady state AUC, steady state volume of distribution, distribution half-life, elimination half-life and accumulation ratio, were estimated from the base popPK model. The distributions of these estimates by study are shown in Figures S9 and S10. Comparisons of these PK features between the two studies adjusted for age, body weight, race, creatinine clearance and dose group using the TMLE approach are shown in Figures 4 and 5. We found that the estimated mean of Vp, steady state volume of distribution, elimination half-life, and accumulation ratio were significantly higher in HVTN 704/HPTN 085 (MSM/TG) than in HVTN 703/HPTN 081 (women): estimated means of Vp 4.86 L and 3.21 L (p < 0.001, adjusted p < 0.001), steady state volume of distribution 7.49 L and 5.58 L (p < 0.001, adjusted p < 0.001), elimination half-life 17.29 and 12.64 days (p=0.005, adjusted p = 0.027) and accumulation ratio 1.11 and 1.04 (p=0.008, adjusted p=0.031) respectively (Table 2, Figure 4, Figure 5).

**Table 2:**
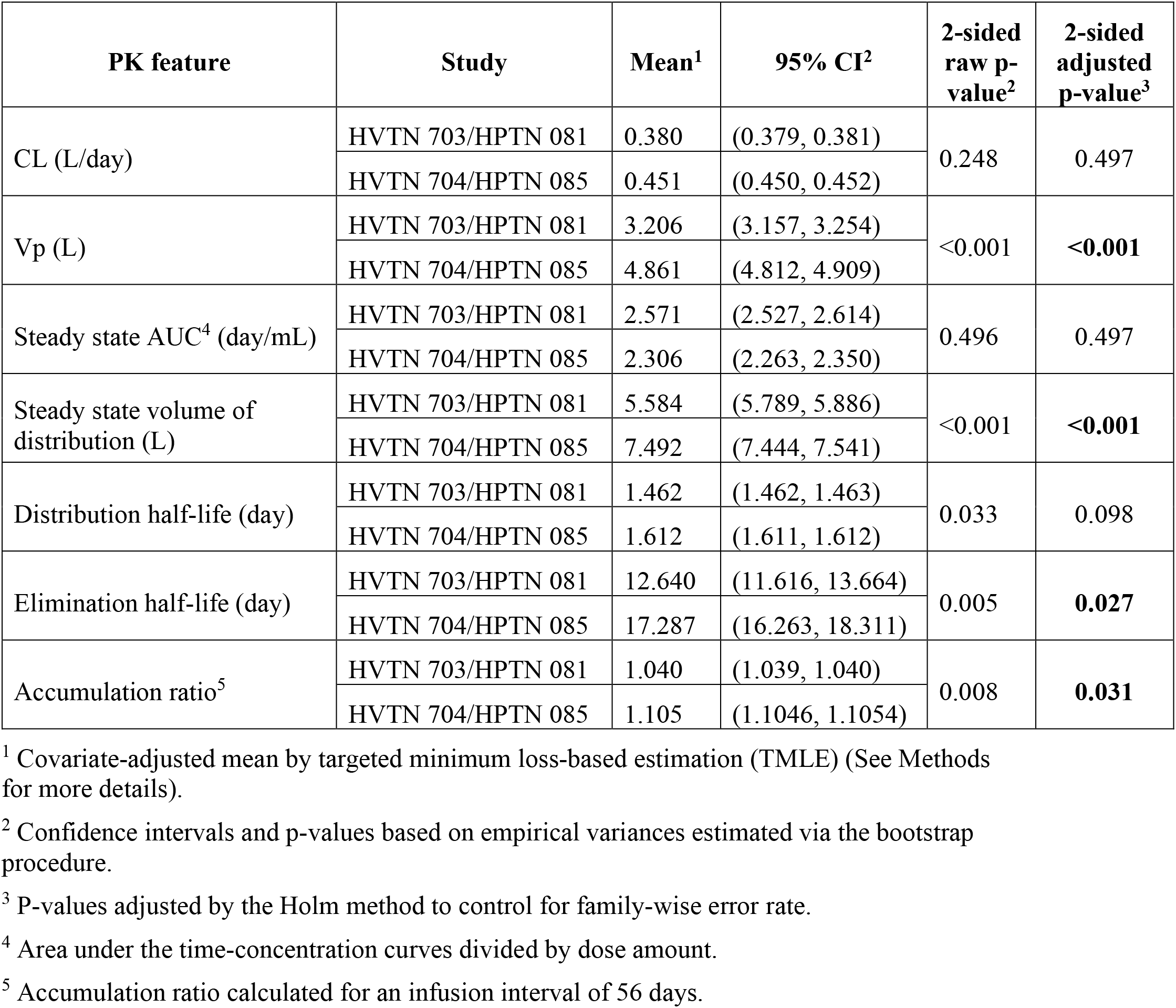
Covariate-adjusted comparisons of PK features between HVTN 703/HPTN 081 (women) and HVTN 704/HPTN 085 (MSM/TG). All comparisons were adjusted for dose, age, body weight, race, and creatinine clearance.

| PK feature | Study | Mean<sup>1</sup> | 95% CI<sup>2</sup> | 2-sided raw p-value<sup>2</sup> | 2-sided adjusted p-value<sup>3</sup> |
| --- | --- | --- | --- | --- | --- |
| CL (L/day) | HVTN 703/HPTN 081 | 0.380 | (0.379, 0.381) | 0.248 | 0.497 |
|  | HVTN 704/HPTN 085 | 0.451 | (0.450, 0.452) |  |  |
| Vp (L) | HVTN 703/HPTN 081 | 3.206 | (3.157, 3.254) | <0.001 | <0.001 |
|  | HVTN 704/HPTN 085 | 4.861 | (4.812, 4.909) |  |  |
| Steady state AUC<sup>4</sup> (day/mL) | HVTN 703/HPTN 081 | 2.571 | (2.527, 2.614) | 0.496 | 0.497 |
|  | HVTN 704/HPTN 085 | 2.306 | (2.263, 2.350) |  |  |
| Steady state volume of distribution (L) | HVTN 703/HPTN 081 | 5.584 | (5.789, 5.886) | <0.001 | <0.001 |
|  | HVTN 704/HPTN 085 | 7.492 | (7.444, 7.541) |  |  |
| Distribution half-life (day) | HVTN 703/HPTN 081 | 1.462 | (1.462, 1.463) | 0.033 | 0.098 |
|  | HVTN 704/HPTN 085 | 1.612 | (1.611, 1.612) |  |  |
| Elimination half-life (day) | HVTN 703/HPTN 081 | 12.640 | (11.616, 13.664) | 0.005 | 0.027 |
|  | HVTN 704/HPTN 085 | 17.287 | (16.263, 18.311) |  |  |
| Accumulation ratio<sup>5</sup> | HVTN 703/HPTN 081 | 1.040 | (1.039, 1.040) | 0.008 | 0.031 |
|  | HVTN 704/HPTN 085 | 1.105 | (1.1046, 1.1054) |  |  |

<sup>1</sup> Covariate-adjusted mean by targeted minimum loss-based estimation (TMLE) (See Methods for more details).
<sup>2</sup> Confidence intervals and p-values based on empirical variances estimated via the bootstrap procedure.
<sup>3</sup> P-values adjusted by the Holm method to control for family-wise error rate.
<sup>4</sup> Area under the time-concentration curves divided by dose amount.
<sup>5</sup> Accumulation ratio calculated for an infusion interval of 56 days.

**Figure 4:**
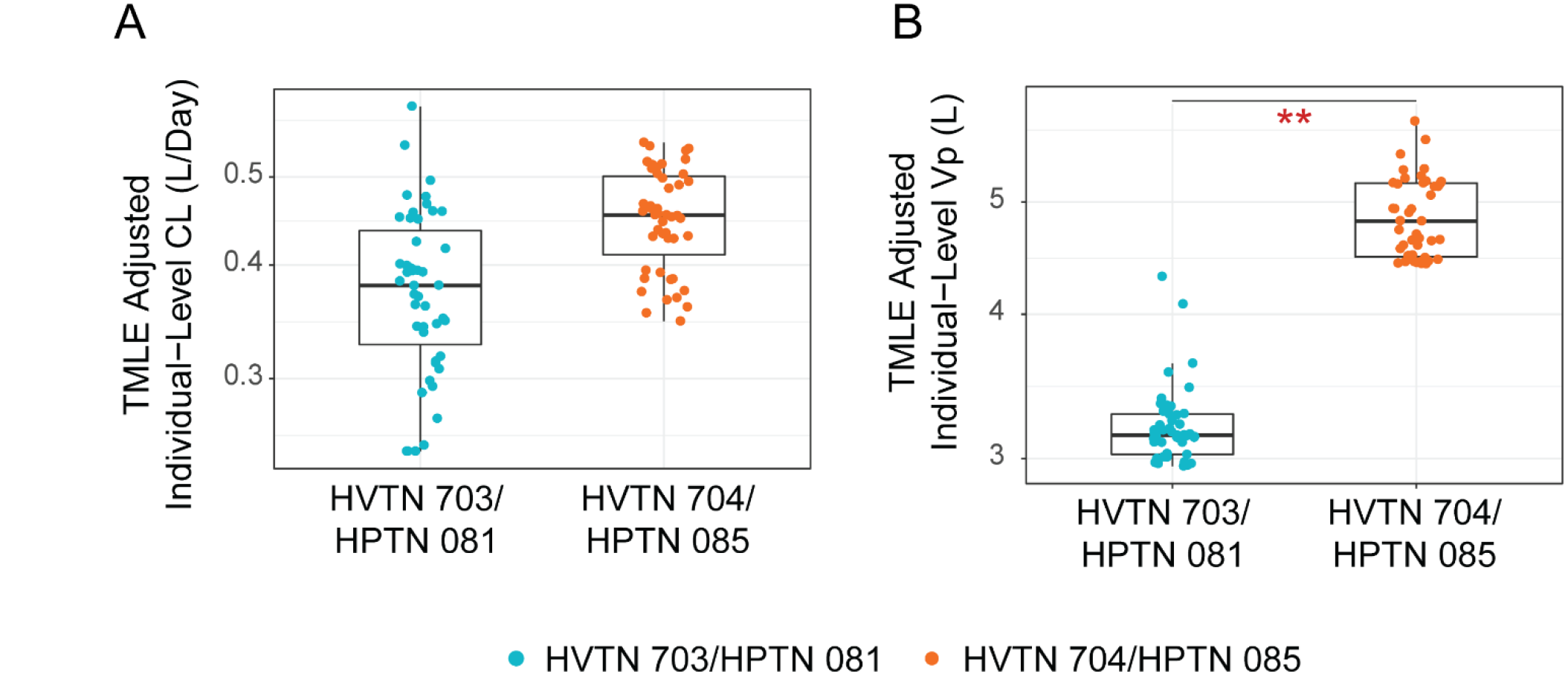
Distributions of covariate-adjusted individual-level PK parameters of VRC01. A) 249 Clearance (CL) and B) volume of the peripheral compartment (Vp). All estimates were adjusted 250 for dose, age, body weight, race, and creatinine clearance via targeted minimum loss-based 251 estimation (TMLE) as presented in Table 2. **, two-sided adjusted p-value < 0.001.

**Figure 5:**
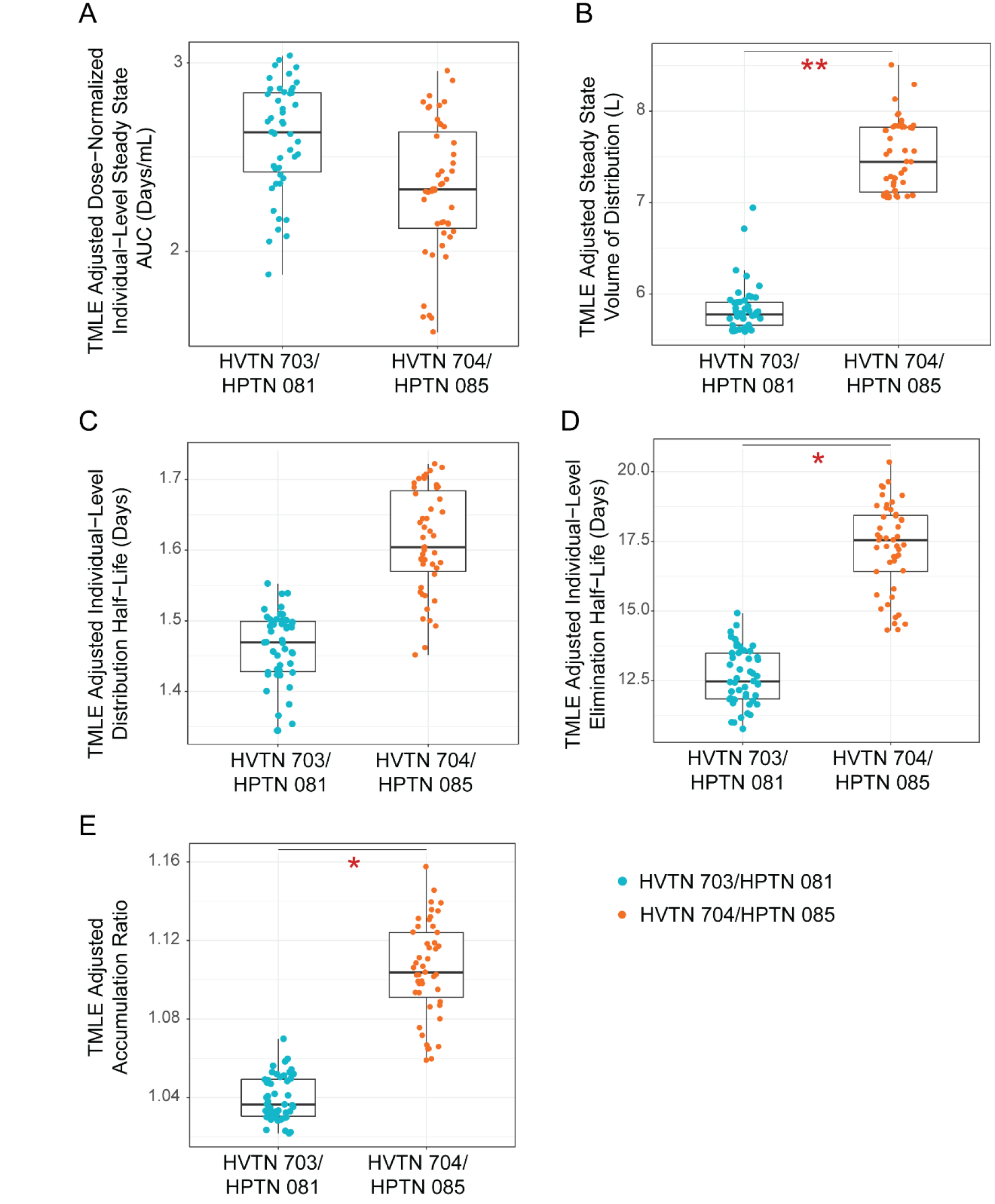
Distributions of covariate-adjusted individual-level PK parameters of VRC01. A) Steady state AUC, B) steady state volume of distribution, C) distribution half-life, D) elimination half-life, and E) accumulation ratio. All estimates were adjusted for dose, age, body weight, race, and creatinine clearance via TMLE as presented in Table 2. *, two-sided adjusted p-value < 0.05; **, two-sided adjusted p-value < 0.001.

## Discussion

This popPK modeling work of VRC01 is the first of its kind to systematically characterize and compare PK of an HIV broadly neutralizing mAb in healthy adults who are at risk of HIV acquisition in two distinct study populations: predominantly black, sub-Saharan African women (AMP HVTN 703/HPTN 081), and predominantly non-black men and transgender persons in the Americas and Switzerland who have sex with men (MSM/TG) (AMP HVTN 704/HPTN 085). Based on VRC01 serum concentration data collected longitudinally over 10 infusions at two different doses from a subset of AMP participants, we constructed a popPK model accounting for baseline participant characteristics, variabilities of PK features across individuals, as well as variabilities across repeated product infusions. We found that participants’ body weight significantly influenced VRC01 clearance in both AMP study populations with a faster clearance for heavier individuals, consistent with findings from previous PK studies of VRC01 in the US and of multiple other mAbs [24]. Mechanisms of underlying PK differences by body weight that should be considered in interpreting these findings include decreased lymph flow rates in obese patients, which may influence the rate and extent of mAb distribution in tissues; increased protein endocytosis and catabolism in underweight patients, which affects clearance; and the correlation of body size with plasma and interstitial fluid volumes, which affect distribution [24].

We identified four PK features of VRC01 (peripheral volume of distribution, steady state volume of distribution, elimination half-life, and accumulation ratio) that were significantly different between the two study cohorts even after adjusting for potential confounding factors including dose, age, race, body weight, and creatinine clearance. This finding suggests that these differences in PK features are likely due to other factors that differ between the two study cohorts, e.g. sex assigned at birth, exposure to pathogens, or genetics. The many cohort differences make it difficult to identify the exact causes of the observed differences in PK features.

There is a paucity of data assessing study population effects on pharmacokinetics of mAbs. Similar PK were observed in American Caucasians and Asian people from Japan and China for evolocumab, a mAb for prevention of hypercholesterolemia-related heart attack and stroke [25]. Likewise, there was no significant difference in genotype frequencies of Fcγ-receptor IIA (receptor-mediated endocytosis via Fcγ receptors may contribute to elimination of some mAbs [19]) between Caucasians and African-Americans [26]. However, to our knowledge, there is no literature comparing PK responses between black Africans and other racial/ethnic groups. As nearly 70% of all people living with HIV globally [27] and over 60% of all new HIV infections are in sub-Saharan Africa [27], there is a particularly pressing need for safe and efficacious HIV prevention and treatment interventions in this region. It is thus a major strength of our study that we characterized PK responses to bnAb infusions in a sub-Saharan African population.

Another difference between the study populations that may have influenced PK features is sex assigned at birth. Although early phase trials evaluating other mAbs for prevention of viral infections, including cytomegalovirus in renal transplant recipients [28] and post-exposure prophylaxis of rabies [29], have not reported any differences in PK responses by sex assigned at birth, this may not be translatable to HIV-1 where sexual acquisition occurs in tissues with significant sex-based physiological differences: vaginal versus rectal. Our study showed that the female assigned at birth participants in HVTN 703/HPTN 081 had a smaller Vp than the predominantly male assigned at birth participants in HVTN 704/HPTN 085. Smaller Vp could be indicative of higher peripheral concentration that may influence efficacy. Further evaluation of VRC01 or similar mAbs for HIV prevention may warrant physiologically based pharmacokinetic (PBPK) modelling to understand PK features at the target tissue-level [30].

We also observed low VRC01 accumulation (≤10%) over the 10 study infusions in both study populations and across both dose groups, although there appeared to be noticeable variabilities of CL and Vp over infusion intervals. Because there was no specific trend in CL and Vp over infusion intervals, these variabilities are more likely due to fluctuation of individual participant characteristics over time, rather than systematic change of the PK features due to repeated dosing. Given the apparent increase in the precision of the inter-individual variability of CL and Vp after incorporating variabilities over infusion intervals, it will be important to account for both types of variability for a more accurate estimation of concentration at time of infection in the future AMP case-control correlates study.

We also presented unique data of the predicted VRC01 neutralization coverage of circulating strains of HIV-1. These plots provide a way to predict the proportion of HIV-1 strains to which trial participants in the geographic areas of each trial may be exposed that would be expected to be neutralized based on the modeled VRC01 serum concentration in each target population. While VRC01 exhibits relatively broad neutralization activity, its neutralization coverage varies across clade, with clade C viruses generally neutralized less well than clade B viruses [31, 32]. This observation was borne out in our predicted coverage plots, where the predicted VRC01 neutralization coverage against the clade C panel was always less than the predicted VRC01 neutralization coverage against the clade B panel, within each dose group, and within the same post-infusion time window (weeks 1-4 vs. weeks 4-8). As serum neutralizing titer against a given exposing virus (calculated by dividing the serum concentration of the bnAb on the day of exposure by the known in vitro titer of that bnAb against the exposing virus) has been shown to be strongly correlated with protection against SHIV acquisition in nonhuman primate challenge studies [33], these results suggest that, while VRC01 serum concentration levels over time are similar across the two trials, neutralization coverage against circulating HIV-1 strains in the specific region may be slightly higher in HVTN 704/HPTN 085 (MSM/TG) compared to HVTN 703/HPTN 081 (women), suggesting slightly higher predicted efficacy of VRC01 in the former study.

We also quantified the greater predicted neutralization coverage in the first 4 weeks post-first infusion compared to in the second 4 weeks. An implication is that the time elapsed from the first infusion until the point of exposure is a major factor that could influence whether the infused VRC01 can provide protection against the circulating strains, and that participants who are exposed in the first 4 weeks post-first infusion are more likely to be protected than participants who are exposed in the second 4 weeks. These results also suggest that the overall VRC01 exposure (i.e. AUC of the time-concentration curve) may not be lower in HVTN 703/HPTN 081 (women) vs. HVTN 704/HPTN 085 (MSM/TG), although other PK features may be.

Our findings are important not only for gaining a preliminary understanding of the PK characteristics of VRC01 in the AMP study populations, which is expected to aid the interpretation of the primary efficacy results (expected in Q4 2020), but also for informing the sampling design of serum concentration and other markers of VRC01 for the AMP correlates study. Specifically, if AMP demonstrates that VRC01 is partially efficacious for the prevention of HIV-1, then the estimated serum VRC01 concentration at the time of HIV-1 acquisition, combined with statistical methods for estimating the day of HIV-1 infection [34], can be used to estimate the association of serum concentration with risk of HIV-1 acquisition, and similarly to estimate the association of neutralizing antibody titer to a panel of HIV-1 strains with risk of HIV-1 acquisition. Such correlates of risk could set a benchmark for the required potency of a vaccine-induced neutralizing antibody response to achieve a high level of protection against HIV infection [21], informing optimal dose-regimen selection for the next-generation of mAbs or mAb combinations with cost-saving implications.

Our study is limited by the lack of data on anti-drug antibodies, which are known contributors to increased elimination of mAbs [35]. We were also unable to assess the effect of albumin concentration, which is inversely associated with mAb clearance [19]. However, since participants were required to be healthy with no malignancies, autoimmune conditions or history of renal or hepatic dysfunction, it is less likely that albumin levels would be a significant contributor to mAb clearance in the AMP trials. Other limitations of our study are that our prediction neutralization coverage analyses are based on the premise that clade-specific HIV-1 sequences retrieved from the CATNAP database represent to some degree the sequences of the HIV-1 viruses to which participants in each trial are exposed. However, it is difficult to ascertain how accurately the HIV-1 sequences in CATNAP represent contemporaneously circulating viruses. For example, a study of recently transmitted clade C viruses documented antigenic drift at VRC01 target sites, and found that the clade C viruses became significantly less sensitive to VRC01-mediated neutralization over the last 20 years [31]. Therefore, it is possible that HVTN 703/HPTN 081 (women) participants are exposed to clade C viruses that have naturally acquired a greater level of resistance to VRC01 than is apparent by using HIV-1 sequences from the CATNAP database. In such a scenario, VRC01 coverage in the HVTN 703/HPTN 081 trial would be expected to be even lower than predicted, potentially leading to decreased prevention efficacy. In addition, for simplicity, we used clade C CATNAP viruses to represent the circulating strains to which HVTN 703/HPTN 081 participants were exposed during the trial, and clade B CATNAP viruses to represent the circulating strains to which HVTN 704/HPTN 085 participants were exposed during the trial. While these clades comprise the majority of circulating strains in the two respective trials, it is worth noting that clades A and D predominate in East Africa, and clades C and F also circulate in South America [36], and these differences would also affect actual VRC01 neutralization coverage.

In conclusion, our results have important implications for the AMP correlates study. First, our results suggest that the sampling of controls for the case-control study should be stratified by study, given the differences observed in Vp and derived PK features. Second, in addition to VRC01 serum concentration and sensitivity to VRC01-mediated neutralization of Env-pseudotyped viruses derived from HIV-1 infected trial participants, the PK features studied here could be evaluated as potential correlates of risk, depending on their a priori biological plausibility and the presence of sufficient inter-individual variability. Third, as we confirmed the impact of body weight on PK parameters, specifically clearance from the central compartment, the correlates analyses can adjust for body weight.

## Data Availability

Upon acceptance, the data underlying the findings of this manuscript will be made publicly available at the public-facing HVTN website (https://atlas.scharp.org/).

## Acknowledgements

We thank the HVTN 703/HPTN 081 and HVTN 704/HPTN 085 trial teams and participants.

## Financial Disclosure Statement

This work was supported by the National Institute of Allergy and Infectious Diseases (NIAID) U.S. Public Health Service Grants UM1 AI068614 [LOC: HIV Vaccine Trials Network] and UM1AI068619 [LOC: HIV Prevention Trials Network], UM1 AI068635 [HVTN SDMC FHCRC] and UM1AI068617 [HPTN SDMC], UM1 AI068618 [HVTN Laboratory Center FHCRC] and UM1AI068613 [HPTN Laboratory Center]. The content of this manuscript is solely the responsibility of the authors and does not necessarily represent the official views of the National Institutes of Health. The funders had no role in study design, data collection and analysis, decision to publish, or preparation of the manuscript.

## Declaration of Interests

The authors of this manuscript have read the journal’s policy and have the following competing interests: LC and PBG are recipients of funding from The National Institute of Allergy and Infectious Diseases of the National Institutes of Health, and this publication is a result of activities funded by the NIAID.

## Methods

### Study Procedures

The AMP trials are being conducted in two distinct study populations: HVTN 703/HPTN 081 (ClinicalTrials.gov #NCT02568215) in sub-Saharan Africa (Botswana, Kenya, Malawi, Mozambique, South Africa, Tanzania, and Zimbabwe) in cisgender women who have sex with men (n=1924), and HVTN 704/HPTN 085 (#NCT02716675) in Brazil, Peru, Switzerland, and the United States in MSM/TG (n=2699). Participants were randomized (1:1:1) to receive ten 8-weekly infusions of 10 mg/kg VRC01, 30 mg/kg VRC01, or placebo (further details of the trial design and statistical considerations are given in [21]). Specimens are collected for HIV diagnosis and biomarker measurements at 25 time-points throughout the trial, including at 5 days after the second infusion (day 61), every 4 weeks from week 0 until week 80, and at weeks 88, 96 and 104.

We analyzed serum concentration data collected at the above time-points from a total of 47 randomly selected VRC01 recipients, 23 from HVTN 703/HPTN 081 (women) and 24 from HVTN 704/HPTN 085 (MSM/TG), with 11 or 12 from each dose group per study (Table 1). Participants were eligible for sampling into the pilot study, irrespective of the number of infusions received and the timing of infusions, if they had remained HIV-1 uninfected until at least week 88, had not permanently discontinued infusions during trial follow-up, and were inferred to have not used PrEP. The HVTN 704/HPTN 085 (MSM/TG) participants were determined to be non-PrEP users based on self-report and dried blood spot data; HVTN 703/HPTN 081(women) were determined to be non-PrEP users based on self-report data given the low frequency of PrEP use in the region (see Text S1 for details). For each sampled participant, serum samples from all available stored sample time points were assayed for VRC01 levels through week 104.

### Serum concentration measurements

Enzyme-linked immunosorbent assay (ELISA) methods were developed to quantify bnAb concentration in human serum [16, 37]. Quantification of VRC01 concentrations in participant serum was performed in 96-well plates on a Beckman Biomek-based automation platform according to the VRC/NVITAL Standard Operating Procedure “5500-Automated ELISA on SCARA Core System.” The VRC01 anti-idiotype, Fab-specific 5C9 monoclonal antibody (manufactured by the Vaccine Research Center, National Institutes of Health) was coated onto Immulon-4HXB microtiter plates overnight at 4^°^C at a concentration of 3.5 μg/mL (concentration is determined for each lot). Plates were then washed and nonspecific binding sites were blocked (10% fetal bovine serum in phosphate-buffered saline) for 2 hours at room temperature. Duplicate serial 3-fold dilutions covering the range of 100 - 24300 of the test sample were incubated for 2 hours at 37^°^C, followed by incubation with horseradish peroxidase-labeled goat anti-human antibodies (1 hour, 37^°^C) and 3,3’,5,5’-tetramethylbenzidine substrate (15 min, room temperature). Color development was stopped by addition of sulfuric acid (stop solution 5% H2SO4), after which the absorbance of each well at 450 nm was measured within 30 minutes using a Molecular Devices Paradigm plate reader. Final sample concentrations were based upon dilution-corrected concentrations estimated from linear regression of a standard curve covering the range of 5 to 125 ng/mL. Concentration values below the limit of quantification (LoQ=1.0 µg/mL) were replaced by 0.5 µg/mL in all calculations. Sensitivity analyses were performed to evaluate the effect of this censoring value on the modeling results. If there were consecutive measurements below the LoQ, only the first one was included in the modeling.

### Population PK (popPK) modeling

PopPK modeling is a powerful approach where drug concentration data from multiple individuals are evaluated simultaneously using a nonlinear mixed-effects model, which incorporates both fixed effects (that are constant) and random effects (that vary across individuals or over time).

#### Structure Model

VRC01 concentrations over time were analyzed using nonlinear mixed effects modeling with the NONMEM software system (Version 7.4, ICON Development Solutions). The stochastic approximation of expectation-maximization (SAEM) algorithm was used for the estimation of model parameters. An open 2-compartment disposition model with first-order elimination from the central compartment was parameterized in terms of clearance from the central compartment in L/day (CL), volume of the central compartment in L (Vc), inter-compartmental distribution clearance in L/day (Q), and volume of the peripheral compartment in L (Vp).

#### Variability popPK model

The statistical model considered three primary sources of variability around the structure population mean model: inter-individual variability (IIV), inter-occasion variability (IOV), and residual variability (RV) remaining after controlling for other sources of variability in the data. IOV was investigated by considering each infusion as an occasion to account for PK parameter changes between infusions due to, for example, changing number of doses or changing participant characteristics over time that may impact the underlying PK process. For both IIV and IOV, an exponential between-individual and between-occasion random effects model was considered such that the distribution of PK parameters is log-normally distributed, but the random effect is normally distributed. Further details are given in Text S1.

Regarding RV, the additive, proportional, and combination proportional + additive residual error models were all considered and compared. Further details are given in Text S1. The percentage coefficient of variation (%CV) of the error terms and the resulting fit of the different error models based on likelihood ratio tests of the objective function value (OFV) (minus twice the log likelihood of the data, with smaller value indicating better fit) were used in determining the final error model. Statistical significance of a hypothesis testing result is noted based on a 2-sided p-value < 0.05.

#### Covariate model

Identification of baseline covariates predictive of PK variability was performed to better understand the sources of observed inter-individual variability. The baseline covariates that were screened for this analysis were pre-defined, including study (HVTN 703/HPTN 081 or HVTN 704/HPTN 085) and dose group (10 mg/kg or 30 mg/kg) as well as demographic variables: age (years), sex assigned at birth (male or female), body weight (kg), race (black or other), body mass index (kg/m^2^); clinical variables: pulse rate (beats/min), respiratory rate (breaths/min), diastolic blood pressure (mmHg), systolic blood pressure (mmHg), temperature (^°^C); and safety lab variables: Cockcroft-Gault creatinine clearance (L/day), erythrocyte mean corpuscular volume (fL), alanine aminotransferase (units/L), hematocrit (%), hemoglobin (g/dL), platelets (10^3^/mm^3^), leukocyte count (10^3^/mm^3^), lymphocyte count (cells/mm^3^), monocyte count (cells/mm^3^), neutrophil count (cells/mm^3^), basophil count (cells/mm^3^), and eosinophil count (cells/mm^3^). These covariates were screened and selected based on their performance in explaining the observed inter-individual variabilities of the PK parameters using a similar model selection procedure as described in Huang et al. [18] where the IOV random effect term was not included (see Text S1). The IOV term was later evaluated in the final popPK model.

### Simulations of serum concentration and prediction of neutralization coverage

Serum concentrations were simulated both over the course of 8 weeks after a single dose and over the course of ten 8-weekly doses for AMP VRC01 recipients based on their body weight and treatment assignment information using the final popPK model without IOV. We graphed predicted neutralization coverage against clade C viruses for HVTN 703/HPTN 081 and clade B viruses for HVTN 704/HPTN 085 using data from the Compile, Analyze and Tally NAb Panels (CATNAP) database [38]; we predicted that an individual’s serum at a specified time point would achieve “neutralization coverage” of a virus if the geometric mean serum concentration of VRC01 was at least 100-fold greater than the viral in vitro inhibitory concentration 50% (IC50) values, as measured in vitro, e.g. via the TZM-bl target cell assay [39]. We based the 100-fold estimate on the nonhuman primate simian human immunodeficiency virus (SHIV)-challenge model, where protection is achieved by CD4 binding-site bnAbs if serum antibody concentrations are approximately 50 to 100-fold higher than the measured IC50 of the challenge virus [14]. We considered viruses to be resistant to neutralization (i.e., neutralization coverage not achieved) if the IC50 was greater than 10 µg/mL.

### Comparison of PK features between groups with covariate-adjustment

Seven individual-level PK features - CL, Vp, steady state volume of distribution, steady state area under the time-concentration curve (AUC), distribution half-life, elimination half-life, and accumulation ratio estimates aggregated over all infusion occasions - were derived from the base popPK model. For comparing non-randomized groups of interest, such as the two AMP trials, to reduce confounding bias the targeted minimum loss-based estimation (TMLE) method [40, 41] was used to estimate the mean of each feature for each group, adjusted for potential predictors of PK variability: age, body weight, race, creatinine clearance and dose group (implemented in the *tmle* R package [42]). TMLE is an alternative to standard linear or nonlinear regression that can have improved robustness and efficiency. All TMLE estimation results of means were averaged over 20 runs with a fixed random seed on top of the leave-one-out cross-validation estimation procedure to ensure stability of the estimates. The set of learning algorithms used by TMLE for estimating the mean outcome conditional on baseline covariates are listed in Text S1. In addition, to account for variability and co-variability of the individual-level estimates for each PK feature due to the fact that they were derived from a common popPK model, a bootstrap procedure based on 250 datasets was used to calculate the empirical variances of the estimates for each group and to derive the 95% confidence interval, as well as to test for a non-zero mean difference between the two groups. The Holm method [43] was used to adjust for multiple comparisons.

## Notes

### Clinical Trial

NCT02568215, NCT02716675

### Author Declarations

SITESIRB/EC BR-Rio de JaneiroInstituto de Pesquisa ClÃnica Evandro Chagas Ethics Committee CH-LausanneCentre Hospitalier Universitaire Saudois Institutional Review Board PE-IquitosImpacta Ethics Committee PE-Lima-BarrancoImpacta Ethics Committee PE-Lima-San MarcosVia Libre Ethics Committee PE-Lima-San MiguelImpacta Ethics Committee PE-Lima-Via LibreVia Libre Ethics Committee US-Atlanta-Hope ClinicFred Hutch Institutional Review Board US-Atlanta-Ponce de LeonFred Hutch Institutional Review Board US-BirminghamFred Hutch Institutional Review Board US-Boston-BrighamPartners Institutional Review Board US-Boston-FenwayFenway Institutional Review Board US-Chapel HillFred Hutch Institutional Review Board US-ClevelandFred Hutch Institutional Review Board US-Los Angeles-Vine StreetUCLA Office for Human Research Participant Protection US-NashvilleVanderbilit Institutional Review Board US-New York-Bronx PreventionFred Hutch Institutional Review Board US-New York-Harlem PreventionFred Hutch Institutional Review Board US-New York-NYBCFred Hutch Institutional Review Board US-New York-Physicians & SurgeonsFred Hutch Institutional Review Board US-NewarkFred Hutch Institutional Review Board US-PhiladelphiaFred Hutch Institutional Review Board US-RochesterFred Hutch Institutional Review Board US-San FranciscoFred Hutch Institutional Review Board US-SeattleFred Hutch Institutional Review Board US-Washington, DCGeorge Washington University Institutional Review Board

## References

1. UNAIDS. Global HIV & AIDS statistics - 2018 fact sheet. Access date Apr 18, 2019 2018. Available from: http://www.unaids.org/en/resources/fact-sheet.

2. Forsythe SS, McGreevey W, Whiteside A, Shah M, Cohen J, Hecht R, Bollinger LA, Kinghorn A. Twenty Years Of Antiretroviral Therapy For People Living With HIV: Global Costs, Health Achievements, Economic Benefits. Health Aff (Millwood). 2019;38(7):1163–72.

3. Desai M, Field N, Grant R, McCormack S. Recent advances in pre-exposure prophylaxis for HIV. BMJ. 2017;359:j5011. PMC6020995.

4. Bekker LG, Alleyne G, Baral S, Cepeda J, Daskalakis D, Dowdy D, Dybul M, Eholie S, Esom K, Garnett G, Grimsrud A, Hakim J, Havlir D, Isbell MT, Johnson L, Kamarulzaman A, Kasaie P, Kazatchkine M, Kilonzo N, Klag M, Klein M, Lewin SR, Luo C, Makofane K, Martin NK, Mayer K, Millett G, Ntusi N, Pace L, Pike C, Piot P, Pozniak A, Quinn TC, Rockstroh J, Ratevosian J, Ryan O, Sippel S, Spire B, Soucat A, Starrs A, Strathdee SA, Thomson N, Vella S, Schechter M, Vickerman P, Weir B, Beyrer C. Advancing global health and strengthening the HIV response in the era of the Sustainable Development Goals: the International AIDS Society-Lancet Commission. Lancet. 2018;392(10144):312–58. PMC6323648.

5. Haynes BF, Burton DR, Mascola JR. Multiple roles for HIV broadly neutralizing antibodies. Sci Transl Med. 2019; 11(516).

6. McCoy LE. The expanding array of HIV broadly neutralizing antibodies. Retrovirology. 2018;15(1):70. PMC6192334.

7. McCoy LE, Burton DR. Identification and specificity of broadly neutralizing antibodies against HIV. Immunol Rev. 2017;275(1):11–20. PMC5299474.

8. Sok D, Burton DR. Recent progress in broadly neutralizing antibodies to HIV. Nat Immunol. 2018;19(11): 1179-88. PMC6440471.

9. Hessell AJ, Malherbe DC, Haigwood NL. Passive and active antibody studies in primates to inform HIV vaccines. Expert Rev Vaccines. 2018;17(2):127–44. PMC6587971.

10. Pegu A, Hessell AJ, Mascola JR, Haigwood NL. Use of broadly neutralizing antibodies for HIV-1 prevention. Immunol Rev. 2017;275(1):296–312. PMC5314445.

11. Pegu A, Yang ZY, Boyington JC, Wu L, Ko SY, Schmidt SD, McKee K, Kong WP, Shi W, Chen X, Todd JP, Letvin NL, Huang J, Nason MC, Hoxie JA, Kwong PD, Connors M, Rao SS, Mascola JR, Nabel GJ. Neutralizing antibodies to HIV-1 envelope protect more effectively in vivo than those to the CD4 receptor. Sci Transl Med. 2014;6(243):243ra88. PMC4562469.

12. Wu X, Yang ZY, Li Y, Hogerkorp CM, Schief WR, Seaman MS, Zhou T, Schmidt SD, Wu L, Xu L, Longo NS, McKee K, O’Dell S, Louder MK, Wycuff DL, Feng Y, Nason M, Doria-Rose N, Connors M, Kwong PD, Roederer M, Wyatt RT, Nabel GJ, Mascola JR. Rational design of envelope identifies broadly neutralizing human monoclonal antibodies to HIV-1. Science. 2010;329(5993):856–61. PMC2965066.

13. Zhou T, Georgiev I, Wu X, Yang ZY, Dai K, Finzi A, Kwon YD, Scheid JF, Shi W, Xu L, Yang Y, Zhu J, Nussenzweig MC, Sodroski J, Shapiro L, Nabel GJ, Mascola JR, Kwong PD. Structural basis for broad and potent neutralization of HIV-1 by antibody VRC01. Science. 2010;329(5993):811–7. PMC2981354.

14. Ko SY, Pegu A, Rudicell RS, Yang ZY, Joyce MG, Chen X, Wang K, Bao S, Kraemer TD, Rath T, Zeng M, Schmidt SD, Todd JP, Penzak SR, Saunders KO, Nason MC, Haase AT, Rao SS, Blumberg RS, Mascola JR, Nabel GJ. Enhanced neonatal Fc receptor function improves protection against primate SHIV infection. Nature. 2014;514(7524):642–5. PMC4433741.

15. Moldt B, Rakasz EG, Schultz N, Chan-Hui PY, Swiderek K, Weisgrau KL, Piaskowski SM, Bergman Z, Watkins DI, Poignard P, Burton DR. Highly potent HIV-specific antibody neutralization in vitro translates into effective protection against mucosal SHIV challenge in vivo. Proc Natl Acad Sci U S A. 2012;109(46):18921–5. PMC3503218.

16. Ledgerwood JE, Coates EE, Yamshchikov G, Saunders JG, Holman L, Enama ME, DeZure A, Lynch RM, Gordon I, Plummer S, Hendel CS, Pegu A, Conan-Cibotti M, Sitar S, Bailer RT, Narpala S, McDermott A, Louder M, O’Dell S, Mohan S, Pandey JP, Schwartz RM, Hu Z, Koup RA, Capparelli E, Mascola JR, Graham BS, Team VRCS. Safety, pharmacokinetics and neutralization of the broadly neutralizing HIV-1 human monoclonal antibody VRC01 in healthy adults. Clin Exp Immunol. 2015;182(3):289–301. PMC4636891.

17. Mayer KH, Seaton KE, Huang Y, Grunenberg N, Isaacs A, Allen M, Ledgerwood JE, Frank I, Sobieszczyk ME, Baden LR, Rodriguez B, Van Tieu H, Tomaras GD, Deal A, Goodman D, Bailer RT, Ferrari G, Jensen R, Hural J, Graham BS, Mascola JR, Corey L, Montefiori DC, Team HP, and the NHIVVTN. Safety, pharmacokinetics, and immunological activities of multiple intravenous or subcutaneous doses of an anti-HIV monoclonal antibody, VRC01, administered to HIV-uninfected adults: Results of a phase 1 randomized trial. PLoS Med. 2017;14(11):e1002435. PMC5685476.

18. Huang Y, Zhang L, Ledgerwood J, Grunenberg N, Bailer R, Isaacs A, Seaton K, Mayer KH, Capparelli E, Corey L, Gilbert PB. Population pharmacokinetics analysis of VRC01, an HIV-1 broadly neutralizing monoclonal antibody, in healthy adults. MAbs. 2017;9(5):792–800. PMC5524155.

19. Ryman JT, Meibohm B. Pharmacokinetics of Monoclonal Antibodies. CPT Pharmacometrics Syst Pharmacol. 2017;6(9):576–88. PMC5613179.

20. Kamath AV. Translational pharmacokinetics and pharmacodynamics of monoclonal antibodies. Drug Discov Today Technol. 2016;21-22:75-83.

21. Gilbert PB, Juraska M, deCamp AC, Karuna S, Edupuganti S, Mgodi N, Donnell DJ, Bentley C, Sista N, Andrew P, Isaacs A, Huang Y, Zhang L, Capparelli E, Kochar N, Wang J, Eshleman SH, Mayer KH, Magaret CA, Hural J, Kublin JG, Gray G, Montefiori DC, Gomez MM, Burns DN, McElrath J, Ledgerwood J, Graham BS, Mascola JR, Cohen M, Corey L. Basis and Statistical Design of the Passive HIV-1 Antibody Mediated Prevention (AMP) Test-of-Concept Efficacy Trials. Stat Commun Infect Dis. 2017;9(1). PMC5714515.

22. Hemelaar J. The origin and diversity of the HIV-1 pandemic. Trends Mol Med. 2012;18(3):182–92.

23. Gilbert PB, Zhang Y, Rudnicki E, Huang Y. Assessing pharmacokinetic marker correlates of outcome, with application to antibody prevention efficacy trials. Stat Med. 2019;38(23):4503–18. PMC6736712.

24. Thomas VA, Balthasar JP. Understanding Inter-Individual Variability in Monoclonal Antibody Disposition. Antibodies (Basel). 2019;8(4). PMC6963779.

25. Wang C, Zheng Q, Zhang M, Lu H. Lack of ethnic differences in the pharmacokinetics and pharmacodynamics of evolocumab between Caucasian and Asian populations. Br J Clin Pharmacol. 2019;85(1): 114-25. PMC6303218.

26. Reilly AF, Norris CF, Surrey S, Bruchak FJ, Rappaport EF, Schwartz E, McKenzie SE. Genetic diversity in human Fc receptor II for immunoglobulin G: Fc gamma receptor IIA ligand-binding polymorphism. Clin Diagn Lab Immunol. 1994;1(6):640–4. PMC368380.

27. AVERT. Global HIV and AIDS statistics. 2018. https://www.avert.org/global-hiv-and-aids-statistics Last update 18 Feb 2020. Accessed 3 May 2020..

28. Ishida JH, Patel A, Mehta AK, Gatault P, McBride JM, Burgess T, Derby MA, Snydman DR, Emu B, Feierbach B, Fouts AE, Maia M, Deng R, Rosenberger CM, Gennaro LA, Striano NS, Liao XC, Tavel JA. Phase 2 Randomized, Double-Blind, Placebo-Controlled Trial of RG7667, a Combination Monoclonal Antibody, for Prevention of Cytomegalovirus Infection in High-Risk Kidney Transplant Recipients. Antimicrob Agents Chemother. 2017;61(2). PMC5278717.

29. Bakker AB, Python C, Kissling CJ, Pandya P, Marissen WE, Brink MF, Lagerwerf F, Worst S, van Corven E, Kostense S, Hartmann K, Weverling GJ, Uytdehaag F, Herzog C, Briggs DJ, Rupprecht CE, Grimaldi R, Goudsmit J. First administration to humans of a monoclonal antibody cocktail against rabies virus: safety, tolerability, and neutralizing activity. Vaccine. 2008;26(47):5922–7.

30. Glassman PM, Balthasar JP. Physiologically-based modeling of monoclonal antibody pharmacokinetics in drug discovery and development. Drug Metab Pharmacokinet. 2019;34(1):3–13. PMC6378116.

31 Rademeyer C, Korber B, Seaman MS, Giorgi EE, Thebus R, Robles A, Sheward DJ, Wagh K, Garrity J, Carey BR, Gao H, Greene KM, Tang H, Bandawe GP, Marais JC, Diphoko TE, Hraber P, Tumba N, Moore PL, Gray GE, Kublin J, McElrath MJ, Vermeulen M, Middelkoop K, Bekker LG, Hoelscher M, Maboko L, Makhema J, Robb ML, Abdool Karim S, Abdool Karim Q, Kim JH, Hahn BH, Gao F, Swanstrom R, Morris L, Montefiori DC, Williamson C. Features of Recently Transmitted HIV-1 Clade C Viruses that Impact Antibody Recognition: Implications for Active and Passive Immunization. PLoS Pathog. 2016;12(7):e1005742. PMC4951126.

32. Gach JS, Quendler H, Tong T, Narayan KM, Du SX, Whalen RG, Binley JM, Forthal DN, Poignard P, Zwick MB. A human antibody to the CD4 binding site of gp120 capable of highly potent but sporadic cross clade neutralization of primary HIV-1. Plos One. 2013;8(8):e72054. PMC3753353.

33. Pegu A, Borate B, Huang Y, Pauthner MG, Hessell AJ, Julg B, Doria-Rose NA, Schmidt SD, Carpp LN, Cully MD, Chen X, Shaw GM, Barouch DH, Haigwood NL, Corey L, Burton DR, Roederer M, Gilbert PB, Mascola JR, Huang Y. A Meta-analysis of Passive Immunization Studies Shows that Serum-Neutralizing Antibody Titer Associates with Protection against SHIV Challenge. Cell Host Microbe. 2019;26(3):336–46 e3. PMC6755677.

34. Rossenkhan R, Rolland M, Labuschagne JPL, Ferreira RC, Magaret CA, Carpp LN, Matsen Iv FA, Huang Y, Rudnicki EE, Zhang Y, Ndabambi N, Logan M, Holzman T, Abrahams MR, Anthony C, Tovanabutra S, Warth C, Botha G, Matten D, Nitayaphan S, Kibuuka H, Sawe FK, Chopera D, Eller LA, Travers S, Robb ML, Williamson C, Gilbert PB, Edlefsen PT. Combining Viral Genetics and Statistical Modeling to Improve HIV-1 Time-of-infection Estimation towards Enhanced Vaccine Efficacy Assessment. Viruses. 2019;11(7). PMC6669737.

35. Chirmule N, Jawa V, Meibohm B. Immunogenicity to therapeutic proteins: impact on PK/PD and efficacy. AAPS J. 2012;14(2):296–302. PMC3326159.

36. Gartner MJ, Roche M, Churchill MJ, Gorry PR, Flynn JK. Understanding the mechanisms driving the spread of subtype C HIV-1. EBioMedicine. 2020;53:102682. PMC7047180.

37. Lynch RM, Boritz E, Coates EE, DeZure A, Madden P, Costner P, Enama ME, Plummer S, Holman L, Hendel CS, Gordon I, Casazza J, Conan-Cibotti M, Migueles SA, Tressler R, Bailer RT, McDermott A, Narpala S, O’Dell S, Wolf G, Lifson JD, Freemire BA, Gorelick RJ, Pandey JP, Mohan S, Chomont N, Fromentin R, Chun TW, Fauci AS, Schwartz RM, Koup RA, Douek DC, Hu Z, Capparelli E, Graham BS, Mascola JR, Ledgerwood JE, Team VRCS. Virologic effects of broadly neutralizing antibody VRC01 administration during chronic HIV-1 infection. Sci Transl Med. 2015;7(319):319ra206.

38. Yoon H, Macke J, West AP, Jr., Foley B, Bjorkman PJ, Korber B, Yusim K. CATNAP: a tool to compile, analyze and tally neutralizing antibody panels. Nucleic Acids Res. 2015;43(W1):W213-9. PMC4489231.

39. Todd CA, Greene KM, Yu X, Ozaki DA, Gao H, Huang Y, Wang M, Li G, Brown R, Wood B, D’Souza MP, Gilbert P, Montefiori DC, Sarzotti-Kelsoe M. Development and implementation of an international proficiency testing program for a neutralizing antibody assay for HIV-1 in TZM-bl cells. J Immunol Methods. 2012;375(1-2):57-67. PMC3332116.

40. Van Der Laan MJ, Rubin D. Targeted maximum likelihood learning. The International Journal of Biostatistics. 2006;2(1).

41. Benkeser D, Carone M, Laan MJV, Gilbert PB. Doubly robust nonparametric inference on the average treatment effect. Biometrika. 2017;104(4):863–80. PMC5793673.

42. Gruber S, van der Laan MJ. tmle: An R package for targeted maximum likelihood estimation. Journal of Statistical Software. 2012;51(13).

43. Holm S. A Simple Sequentially Rejective Multiple Test Procedure. Scand J Stat. 1979;6(2):65–70.

